# Concordance of “rapid” serological tests and IgG and IgM chemiluminescence for SARS-COV-2

**DOI:** 10.1101/2020.06.01.20114884

**Authors:** K Sáenz-Flor, L y Sanatafé

**Affiliations:** School of Medicine. Central University Ecuador; Synlab Solutions in Diagnostics Ecuador

## Abstract

**Background:** The COVID-19 serological tests for IgG and IgM have been developed with several methodologies: Immunoenzymatic Assay (ELISA), Chemiluminescence, Electro Chemiluminescence, Fluorescent Lateral Flow Immunoassays and Immunochromatography. None of these tests should be used for the diagnosis or population screening of the disease, considering that the antibodies appear only on the 8th – 14th day of the disease onset. The present study evaluates a sample of immunofluorescent and immunochromatographic rapid tests to show their agreement in relation to Chemiluminescence.

**Methods:** A diagnostic test evaluation assay was performed to establish the performance of five “rapid” tests (4 immunochromatographic and 1 immunofluorescent tests) for IgG and IgM serology for SARS-CoV-2 using a panel of 30 serum samples from patients received in the laboratory analysis routine. For the evaluation of clinical performance, the qualitative results of the “rapid” tests were compared against those obtained by chemiluminescence, dichotomized as positives (≥ 10 AU / mL) or negative (<10 UA / mL).

**Findings:** The best agreement is seen in the immunofluorescent assay, for the IgG contrast, with a particularly good kappa index (0.85), without positive disagreements and a negative disagreement of about 15%. In the immunochromatographic methods Kappa index was 0.61 at best, with disagreements in negative findings of ≈35% and in positive cases of up to ≈70%.

The IgM concordance behavior, on the other hand, reflects a weak to moderate Kappa concordance value (Kappa 0.2 to 0.6), with negative disagreements reaching up to 55% and positives of up to 84%, without any evaluated test reaching Kappa performance equal to or greater than 0.8.

**Interpretation:** Serological studies should be used in the clinical and epidemiological context and of other diagnostic tests. Given the high demand and supply in the market of “rapid serological tests”, its evaluation against panels of serologically positive or negative samples established by Chemiluminescence or Electro chemiluminescence is essential to authorize its extensive use in populations

**Funding:** None

## Introduction

In early 2020, the World Health Organization declared a global health emergency due to the outbreak of a new coronavirus, initially called 2019-nCOV and later called SARS-CoV-2, causing a severe acute respiratory syndrome, Corona Virus Disease 2019 (COVID-19).(1,2).

It is a zoonotic coronavirus that causes, in its severe forms, an acute respiratory distress syndrome that in most cases can occur with mild symptoms. (3,4). As of May 12, an overall case fatality of 6.9% (283,153 deaths / 4,088,848 confirmed cases) is reported, in the Americas 5.9% (104,549 deaths / 1,774,371 confirmed cases), and for Ecuador 7.3% (2,145 deaths / 29509 confirmed cases) according to the official bulletins of the World Health Organization (5)

The COVID-19 serological tests for IgG and IgM have been developed with several methodologies: Immunoenzymatic Assay (ELISA), Chemiluminescence, Electro Chemiluminescence, Fluorescent Lateral Flow Immunoassays and Immunochromatography. Each one has a different and variable performance in relation to the clinical moment in which they are used, as well as whether they were developed to detect S (Spike) or N (Nucleocapsid) antigens, being the former apparently more sensitive.(6) None of these tests should be used for the diagnosis or population screening of the disease, considering that the antibodies appear only on the 8th – 14th day of the disease onset. (7).

Laboratory tests that detect antibodies to SARS-CoV-2, including rapid immunodiagnostic tests, need validation to determine their accuracy and reliability, since there is a double risk. The first one is to falsely label people who have been infected as sero-negative, and the second one is that people who have not been infected are falsely labeled as sero-positive; both errors have serious consequences. In addition to this, it is necessary to ensure that these tests distinguish between infections caused by SARS-CoV-2 and those caused by other human coronaviruses (cross reaction).(8)

The rapid development associated with the market urgency has meant that these tests do not have a solid clinical validation, which makes them show divergences in sensitivity and specificity in their use. (6)

In Ecuador, 102 rapid tests of various brands and methodologies have been authorized by the Health Authorities so far. (9)

The present study evaluates a sample of immunofluorescent and immunochromatographic rapid tests to show their agreement in relation to Chemiluminescence.

## METHODOLOGY

A diagnostic test evaluation assay was performed to establish the performance of five “rapid” tests (4 immunochromatographic and 1 immunofluorescent tests) for IgG and IgM serology for SARS-CoV-2.

The evaluation was performed by comparing to a YHLO_iFLASH_1800 ™ Chemiluminescent analyzer, in a medical laboratory with ISO 9001: 2015 certification and with ISO 15189: 2012 accreditation, in the city of Quito. This device was previously verified according to CLSI EP15 A3(10), with a CV of 3.3% for repeatability and 5.4% for intermediate precision, within manufacturer’s recommendations (<10%)(11,12). Additional performance characteristics are shown in Table 1.

**Table 1.** **Performance characteristics. YHLO- iFLASH_1800 ™**.(11–14)

| Characteristic | SARS-CoV-2 IgG | SARS-CoV-2 IgM |
| --- | --- | --- |
| Detection limit | 6 AU / mL | 5 AU / mL |
| Analytical specificity | No interference with CMV, M Pneumonia, Chlamydia pn, Rubella, EB VCA, EBNA IgG |  |
| Diagnostic sensitivity * | 97.3% | 86.1% |
| Diagnostic specificity * | 96.3% | 92.2 % |

\* Samples from patients with positive PCR (n = 331).
Taken from: SARS-CoV-2 IgG iFLASH v3.0 / SARS-CoV-2 IgM iFLASH v3.0

The results obtained from the rapid tests were compared against reports from the chemiluminescent analyzer, using a panel of 30 serum samples from patients received in the laboratory analysis routine. The samples were taken by venepuncture in the anterior side of the elbow, using a multiple-extraction device (Vacuette®), with Vacuette® 21G needle and BD Vacuntainer® tube, with clot activator and gel separator, for serum collection. Samples were mixed by gentle inversion(15). The tubes were vertically placed on a rack for 30 minutes at room temperature and then were centrifuged at 2000g for 10 minutes. All samples were free of hemolysis, lipemia, and jaundice.

The signals obtained by the automated analyzer for IgG and IgM were classified as Positive where > 10 AU / mL (13/30) for IgG and (6/30) for IgM. (11,12)

For the evaluation of clinical performance, the qualitative results of the “rapid” tests were compared against those obtained by chemiluminescence, dichotomized as positives (≥ 10 AU / mL) or negative (<10 UA / mL). A total of 5 “rapid” tests were compared, 4 of them immunochromatographic and 1 fluorescent immunoassay.

As there is no state-of-the-art “reference standard”, this study does not state sensitivity or specificity, nor predictive values, but only the demonstration of agreement in defining the subjects with the dichotomous criterion (positive or negative), by calculating complex repeatability (global percentage of agreements), percentage of positive agreements and of negative agreements, together with their corresponding 95% confidence interval and complemented with Cohen’s Kappa test as a measure of agreement that discriminates between agreements by chance.(16).

With the obtained data, a Microsoft Excel database was created for subsequent refining and analysis by means of the JASP® software. The luminescent chemo signals detected in the samples were expressed in means and standard deviations, while the concordance were provided in percentages, accompanied by their corresponding confidence interval and through the Cohen’s Kappa index.(16).

The Research Committee of the Faculty of Medical Science of the Central University of Ecuador review the study and conclude that ethical approval was not required, because the samples of patient are used in the context of clinical diagnoses, and do not exist intervention that affect at clinical decision over patients or their integrity.

## RESULTS

The characterization in AU / mL for IgG and IgM (AU / mL) for the positive and negative samples is shown in Table 2.

**Table 2.** **Characterization (AU / mL) for positive and negative samples IgG and IgM for SARS-COv-2.**

**IgG and IgM for SARS-CoV-2.**
| Indicators | IgG (AU / mL) |  | IgM (AU / mL) |  |
| --- | --- | --- | --- | --- |
|  | + | - | + | - |
| **n** | 13 | 17 | 13 | 17 |
| **Average** | 51.98 | 2.899 | 215.2 | 1.451 |
| **Median** | 53.07 | 1.470 | 8.380 | 1.160 |
| **Standard deviation** | 19.87 | 2.924 | 456.1 | 1.166 |
| **2.5<sup>th</sup> percentile** | 10.23 | 0.3300 | 1.120 | 0.5200 |
| **97.5<sup>th</sup> percentile** | 86.49 | 9.030 | 1407 | 5.580 |

Table 3 summarizes the concordance findings of the diagnostic tests evaluated (n = 5) in reference to Chemoluminescence.

**Table 3.** **Performance Evaluation, “Rapid” Serological Assays using Immunochromatographic and Immunofluorescent methods versus Chemiluminescent Serology.**

| Maker | Type | Measurement principle | Comparison n * | MANUFACTURER |  | IgG |  |  |  | IgM |  |  |  |
| --- | --- | --- | --- | --- | --- | --- | --- | --- | --- | --- | --- | --- | --- |
|  |  |  |  | Diagnostic sensitivity | Diagnostic Specificity | Complex Repeatability % [95% CI] | Kappa | Positive Agreements % [IC<sub>95%</sub>] | Negative Agreements % [IC<sub>95%</sub>] | Complex Repeatability | Kappa | Positive Agreements % [IC<sub>95%</sub>] | Negative Agreements % [IC<sub>95%</sub>] |
| Wondfo Biotech Co | Total Antibodies | Fluorescent Immunoassay (PCT) | 27 | NR | NR | 92.6 [76.6 - 97.9] | 0.85 | 100 [Nc] | 85.7 [63.4 - 92.7] | 66.7 [47.8 - 81.4] | 0.37 | 100 [Nc] | 57.1 [38.1 - 74.0] |
| Vazyme Medical Technology Co. Ltd | IgG - IgM | Immunochromatography | 30 | NR | NR | 80 [62.7 - 90.5] | 0.57 | 53.8 [88.5 - 93.0] | 100 [Nc] | 86.7 [70.3 - 94.7] | 0.63 | 83.3 [50.5 - 88.5] | 87.5 [70.8 - 93.8] |
| Xiamen AmonMed Biotechnology Co., Ltd | IgG - IgM | Immunochromatography | 30 | NR * | NR * | 70 [52.1 - 83.3] | 0.33 | 30.8 [15.5 - 91.8] | 100 [Nc] | 83.3 [66.4 - 92.7] | 0.24 | 16.7 [9.8 - 88.5] | 100 [Nc] |
| Shenzhen Rongjin Technology Co. Ltd | IgG - IgM | Immunochromatography | 27 | 93.30% | 96.60% | 74.1 [55.3 - 86.8] | 0.49 | 83.3 [58.9 - 91.8] | 66.7 [44.1 - 82.4] | 66.7 [47.8 - 81.4] | 0.32 | 83.3 [50.5 - 88.5] | 61.9 [42.4 - 77.7] |
| Wondfo Biotech Co | Total Antibodies | Immunochromatography | 30 | 86.43% | 99.57% | 80 [62.7 - 90.5] | 0.61 | 100 [Nc] | 64.7 [43.3 - 80.7] | 56.7 [39.2 - 72.6] | 0.25 | 100 [Nc] | 45.8 [29.2 - 63.7] |

Nc = Not calculable / NR = Not reported / \* Number 27 is associated with a limitation in the number of tests available to run the evaluation

## DISCUSSION

Given the epidemiological urgency and the growing demand for tests that could contribute to the management of the SARS-CoV pandemic, there has been a high demand for diagnostic tests, including immunoassays, which have had rapid development and commercialization with limited validation. in clinical samples (17)

Given the important penetration of rapid tests, mostly immunochromatographic in Ecuador and which have received marketing authorization by the Ministry of Public Health (16), the present study evaluated the concordance of these tests with a luminescent chemoimmunological analysis, using patients’ samples received at the laboratory for seroprevalence evaluations.

The best agreement (both positive and negative agreements) is seen in the immunofluorescent assay, for the IgG contrast, with a particularly good kappa index (0.85), without positive disagreements and a negative disagreement of about 15%. This contrasts with immunochromatographic methods where the Kappa index was 0.61 at best, with disagreements in negative findings of ≈35% and in positive cases of up to ≈70%.

The variations found may be due to the type of antigens used for the development of the assay. Apparently, according to several publications, if they are oriented to nucleocapsid antigens they would seem to be more sensitive, but if they are oriented to the host binding protein (RBD-S) they would be more specific (18)

The great variation in agreement percentages found should draw attention to the problems of diagnostic certainty of serological tests associated with cross-reactions with other coronaviruses. Added to this is the low proportion of negative agreements and their impact on an erroneous screening of a subject for epidemiological surveillance, which may overestimate the population rate considered as immune, when in fact it is not.(6)

So far, most of these studies show that people who have recovered from an infection have antibodies to the virus. However, some of these persons have very low levels of neutralizing antibodies in their blood. Therefore, until now, it has not been evaluated whether the presence of detectable antibodies against SARS-CoV-2 confers immunity to a subsequent infection.(8)

Serological studies should be used in the clinical and epidemiological context and of other diagnostic tests. (8,19). Given the high demand and supply in the market of “rapid serological tests”, its evaluation against panels of serologically positive or negative samples established by Chemiluminescence or Electro chemiluminescence is essential to authorize its extensive use in populations.

If use is required, it is recommended that they be carried out by trained technical-operational personnel and under the supervision of professionals in laboratory medicine.

## Data Availability

The data that support the findings of this study are available from the corresponding author, [Saenz-Flor,K], upon reasonable request.

